# Spatial and temporal transmission dynamics of respiratory syncytial virus in New Zealand before and after the COVID-19 pandemic

**DOI:** 10.1101/2024.07.15.24310412

**Authors:** Lauren Jelley, Jordan Douglas, Meaghan O’Neill, Klarysse Berquist, Ana Claasen, Jing Wang, Srushti Utekar, Helen Johnston, Bocacao Judy, Margot Allais, Joep de Ligt, Chor Ee Tan, Ruth Seeds, Tim Wood, Nayyereh Aminisani, Tineke Jennings, David Welch, Nikki Turner, Peter McIntyre, Tony Dowell, Adrian Trenholme, Cass Byrnes, The SHIVERS investigation team, Richard Webby, Nigel French, David Winter, Q. Sue Huang, Jemma L Geoghegan

## Abstract

Human respiratory syncytial virus (RSV) is a major cause of acute respiratory infection. In 2020, RSV was effectively eliminated from the community in New Zealand due to non-pharmaceutical interventions (NPI) used to control the spread of COVID-19. However, in April 2021, following a brief quarantine-free travel agreement with Australia, there was a large-scale nationwide outbreak of RSV that led to reported cases more than five times higher, and hospitalisations more than three times higher, than the typical seasonal pattern. In this study, we generated 1,471 viral genomes of both RSV-A and RSV-B sampled between 2015 and 2022 from across New Zealand. Using a phylodynamics approach, we used these data to better understand RSV transmission patterns in New Zealand prior to 2020, and how RSV became re-established in the community following the relaxation of COVID-19 restrictions. We found that in 2021, there was a large epidemic of RSV in New Zealand that affected a broader age group range compared to the usual pattern of RSV infections. This epidemic was due to an increase in RSV importations, leading to several large genomic clusters of both RSV-A ON1 and RSV-B BA9 genotypes in New Zealand. However, while a number of importations were detected, there was also a major reduction in RSV genetic diversity compared to pre-pandemic seasonal outbreaks. These genomic clusters were temporally associated with the increase of migration in 2021 due to quarantine-free travel from Australia at the time. The closest genetic relatives to the New Zealand RSV genomes, when sampled, were viral genomes sampled in Australia during a large, off-season summer outbreak several months prior, rather than cryptic lineages that were sustained but not detected in New Zealand. These data reveal the impact of NPI used during the COVID-19 pandemic on other respiratory infections and highlight the important insights that can be gained from viral genomes.

## Introduction

Respiratory syncytial virus (RSV) is a major cause of acute respiratory infection mainly affecting children, the elderly, and those with comorbidities, resulting in a considerable hospitalisation burden among children in particular^1,2^. This RNA virus is part of the O*rthopneumoviruses*, a genus of viruses in the *Pneumoviradae* family that generally cause upper respiratory tract infections in humans and some other vertebrates^3^. RSV is known to cause severe disease in children, particularly infants^4^, and in older adults. In 2019, for instance, there were an estimated 33 million RSV-associated acute respiratory infections globally, and over 100,000 deaths in children under five years old^5^. While serological studies have found that RSV infection is common in infants^4^, the virus induces poor immunity and thus reinfection can be frequent throughout life^6^. RSV infection is less severe in adults, typically causing upper respiratory infections with mild to moderate symptoms^7^. However, in the elderly, RSV can be more severe, and is reported to have morbidity and mortality rates comparable with influenza virus^8^.

The global circulation of RSV is characterised by a distinct seasonality^9-12^. In New Zealand, a geographically isolated country in the southern hemisphere with a temperate climate, RSV cases usually increase in autumn (i.e. at the end of April) and peak in mid-winter (late June)^13^. However, in 2020, the regular circulation of RSV was disrupted due to the introduction of non-pharmaceutical interventions (NPI), the most stringent of which was the closure of international borders, used to curb the spread of SARS-CoV-2^14^. These NPI were effective in eliminating the circulation and re-introduction of SARS-CoV-2 in New Zealand^15,16^, and consequently appeared to eliminate the introduction and spread of other respiratory viruses too^14^. During the winter of 2020, reported cases of RSV were reduced by 98% when compared to the average annual cases recorded from 2015 to 2019^14^. Indeed, compared to annual averages, New Zealand also saw reductions in cases of influenza virus (99.9%), metapneumovirus (92%), enterovirus (82%), adenovirus (81%), parainfluenza virus (80%), and rhinovirus (75%)^14^. A decrease in the circulation of RSV during 2020 was also observed in other countries that used NPI including in the USA^17^, Australia^11^, China^18^, Singapore^19^, South Africa^20^, UK, and Ireland^21^.

In April 2021, following over a year of restricted travel into New Zealand, a brief quarantine-free travel agreement was initiated between New Zealand, Australia, and other countries in the South Pacific^22^. This relaxation of travel restrictions coincided with an increase in the circulation of respiratory viruses in New Zealand^23^. Specifically, New Zealand experienced reported RSV cases five times higher, and hospitalisations three times higher, than those typically seen in the pre-pandemic phase^24^. Australia experienced similar epidemic surges of RSV following the relaxation of COVID-19 restrictions leading to extensive off-season outbreaks and hospitalisations during the summer of 2020 from cryptically circulating lineages^11^. By August 2021, New Zealand had suspended quarantine free travel and once again closed the border with Australia. New Zealand then entered its second nationwide lockdown in order to control the spread of the highly-transmissible SARS-CoV-2 Delta variant^25^. This in turn was met with a considerable decline in the number of reported RSV cases^26^. Beginning early 2022, the New Zealand border became unrestricted, and with the increased migration of people into the country there has been widespread community transmission of respiratory viruses once again, including influenza virus, RSV and the SARS-CoV-2 Omicron variant^26,27^.

Viral genomic data offers valuable insights into the pattern of viral evolution, and therefore enhances our understanding of outbreak epidemiology by providing a high level of detail and context to transmission patterns. When combined with geographic and population information, these data can reveal pathways of viral spread that can be used to inform effective national and local interventions^16,28^. Temporal and geographic information can be integrated into phylodynamic analyses to detect the spread of a pathogen in time and space, develop predictions about an outbreak’s trajectory, and identify clusters of similar sequences as an indication of infection sources and superspreading^28-30^. Herein, we describe the first genome-scale analysis of RSV in New Zealand spanning eight years from 2015 to 2022. This time frame incorporates several significant time points, including the New Zealand border closure in early 2020, nationwide lockdowns during the COVID-19 pandemic, the quarantine-free travel period with Australia and the South Pacific in 2021, and later, the easing of travel restrictions as New Zealand reconnected with the rest of the world in 2022. In this study we investigate the typical patterns and dynamics of RSV transmission in New Zealand prior to 2020, and how RSV became re-established in the community following its elimination.

## Materials & Methods

### Ethics

The New Zealand Northern A Health and Disability Ethics Committee approved the Southern Hemisphere Influenza Vaccine Effectiveness Research and Surveillance (SHIVERS) study (NTX/11/11/102). Ethics approval was not required for the primary care), laboratory, or hospital-based surveillance platforms as they were conducted in accordance with the Public Health Act 1956.

### Sample collection

Viral samples were obtained from several surveillance and research platforms, each serving a different purpose. Community-based samples were obtained from general practitioners (GPs) across New Zealand from patients with influenza-like illness (ILI), for which the surveillance period typically began in late April and ended in late September. Hospital-based samples were obtained from patients with severe acute respiratory infection (SARI) from two major Auckland based hospitals year round.

Laboratory-based surveillance samples were referred to the Institute for Environmental Science and Research (ESR) from both community and hospital diagnostic laboratories throughout the year in several major centres: Auckland, Bay of Plenty, Waikato, Hawkes Bay, Wellington, Nelson, Christchurch and Dunedin. Samples were also collected and tested at ESR for the Southern Hemisphere Influenza Vaccine Effectiveness Research and Surveillance (SHIVERS) study. This study included three Wellington-based community cohorts, where samples were obtained from participants that experienced acute respiratory infection (ARI). The surveillance period for these cohorts matched that of the community-based surveillance. In 2021, a fifth iteration of SHIVERS commenced in which samples from diagnostic and hospital laboratories throughout New Zealand referred positive RSV samples to ESR for further testing and analysis. Collection dates of the genomes used in this study were collected from July 2015 up to, and including, October 2022.

### Viral RNA extraction and qRT-PCR

Nasopharyngeal samples were received by the Clinical Virology Department at ESR from 2015 to 2022. Viral RNA was extracted using the Thermo Fisher Scientific™ MagMax Viral/Pathogen Nucleic Acid Isolation Kit (A48310) as per manufacturer’s instructions on either the ZiXpress or the Kingfisher Flex automated extraction instruments.An RSV singleplex qRT-PCR was performed on RNA using primer and probes synthesised by biosearch using sequences published by Kim et al^31^, coupled with the AgPath-ID™ One step RT-PCR buffer and enzyme (4387424). Positive RSV RNA was then genotyped (RSV-A, RSV-B) using primers and probes synthesised by biosearch using sequences published by Hu et al^32^, coupled with the AgPath-ID™ One step RT-PCR buffer and enzyme (4387424). Samples collected between 2015 and 2021 with a cycle threshold (Ct.) below 25, as well as samples from 2022 with a Ct. below 30, were referred to the genomic sequencing department at ESR.

### Generating viral genomes from RSV samples

In brief, sequencing was performed on the Illumina Nextseq platform using the respiratory virus oligo panel v2 (20044311), RNA prep enrichment kit (20040537) and the 500/550 midoutput kit (20024905) and used as per manufacturer’s instructions. Consensus based assembly was performed using a standardised pipeline, based on the Seattle Flu assembly pipeline, and modified for use by ESR, (https://github.com/seattleflu/assembly). Two additional references were used for consensus calling: EPI_ISL_2543807 and EPI_ISL_2543850, available from GISAID^33^. Consensus viral genomes were generated, and subject to quality testing and those with fewer than 50% ambiguities were selected for further analysis. The genomes generated are available on GISAID under two blocks of consecutive accession numbers (EPI_ISL_16959469 - EPI_ISL_16960152 and EPI_ISL_19206151 - EPI_ISL_19207488).

### Genotyping RSV genomes

Reference G glycoprotein gene (G-gene) sequences for each known genotype for both RSV-A and RSV-B were obtained from GenBank (see Supplementary Table 1 for a list of accession numbers and their genotypes). G-gene sequences were first annotated and extracted from the New Zealand genomes using Geneious Prime 2022.1.1 (https://www.geneious.com) and aligned with the reference sequences using MAFFT (v7)^34^ using the FFT-NS-2 algorithm. A maximum likelihood phylogenetic tree for each of RSV-A and RSV-B was estimated using IQ-TREE (v 1.6.8)^35^, using the Hasegawa-Kishino-Yano nucleotide substitution model with a gamma distributed rate variation among sites (HKY+Γ)^36^ (determined to be the best fit model by ModelFinder)^37^. Using the topology of the reference sequences, the genotypes of the New Zealand RSV G-genes were determined. Genotyping across whole genome sequences was also performed using LABEL^38^ based on genotypes proposed by Chen et al^39^ following alignment with MAFFT (v7)^34^.

**Table 1.** Summary of estimated clock rates and RSV migration events between New Zealand and the rest of the world (BEAST2). Each replicate contains a different subsample of genomes. Mean estimates (and 95% credible intervals) are rounded to 3 significant figures. MCMC chains that did not converge (replicates RSV-B 306) are omitted from the table. *These replicates are shown in Figure 4.

|  | Rep | Clock rate (subs/site/year) | Number of introductions | Number of exports |
| --- | --- | --- | --- | --- |
| **RSV-A** | 1* | $8.79 \times 10^{-4}$ ($8.40 \times 10^{-4}$, $9.13 \times 10^{-4}$) | 85.3 [83, 88] | 2.94 [1, 5] |
| | 2 | $8.31 \times 10^{-4}$ ($7.95 \times 10^{-4}$, $8.66 \times 10^{-4}$) | 81.9 [80, 84] | 3.93 [3, 5] |
| | 3 | $8.25 \times 10^{-4}$ ($7.90 \times 10^{-4}$, $8.59 \times 10^{-4}$) | 89.4 [84, 93] | 2.15 [1, 5] |
| | 4 | $8.68 \times 10^{-4}$ ($8.29 \times 10^{-4}$, $9.04 \times 10^{-4}$) | 83.4 [82, 85] | 3.11 [2, 4] |
| | 5 | $8.52 \times 10^{-4}$ ($8.19 \times 10^{-4}$, $8.87 \times 10^{-4}$) | 84.6 [82, 87] | 3.71 [2, 6] |
| | 6 | $8.68 \times 10^{-4}$ ($8.32 \times 10^{-4}$, $9.04 \times 10^{-4}$) | 80.2 [78, 82] | 4.16 [3, 6] |
| **RSV-B** | 1* | $1.04 \times 10^{-3}$ ($1.00 \times 10^{-3}$, $1.09 \times 10^{-3}$) | 100 [97, 104] | 0.86 [0, 3] |
| | 2 | $1.00 \times 10^{-3}$ ($9.64 \times 10^{-4}$, $1.05 \times 10^{-3}$) | 101 [97, 104] | 0.57 [0, 1] |

### Inferring genomic diversity of RSV using phylogenomic analysis

Newly acquired New Zealand RSV genomes were aligned with globally-sourced RSV genomes with MAFFT (v7)^34^ using the FFT-NS-2 algorithm (see Supplementary Table 2 for global genome accession numbers). These global genomes (3,487 RSV-A and 3,002 RSV-B) were sampled uniformly at random from ∼50,000 international sequences on GISAID^33^ between 2015 and 2022 (as of March 2024). A maximum likelihood phylogenetic tree was estimated using IQ-TREE (v 1.6.8)^35^, using the HKY+Γ^36^ nucleotide substitution model. We integrated the in-build Least Squares Dating (LSD2) method to estimate a time-scaled phylogenetic tree using the day of sampling. Phylogenetic branch support was assessed using the ultrafast bootstrap^40^ approximation. We used TempEst (v 1.5.3)^41^ to analyse the root-to-tip genetic regression against sampling dates to investigate the temporal signal of RSV-A and RSV-B genomes.

### Estimating introductions of RSV using Bayesian phylodynamic analysis

To infer the migration dynamics of RSV into New Zealand over time, we treated geographic location as a discrete trait with two demes where genomes were either sampled as being ‘local’ (i.e. sampled from New Zealand) or ‘global’ (i.e. sampled from the rest of the world). Due to the size of the dataset, local and global RSV-A and RSV-B genomes were randomly split into six subsamples, making sure that near equal numbers of genomes were selected for each deme for each year. For each of RSV-A and RSV-B, each subsample comprised approximately 400 local and 400 global genomes. To correct for geographical biases on GISAID, global genomes were subsampled by iteratively sampling a country and then a genome from that country. Each subsample was aligned using MAFFT (v7)^34^ and subject to Bayesian phylodynamic analysis using BEAST2 (v2.7)^42^ with the MASCOT structured coalescent tree prior^43^. We again used the HKY+Γ^36^ nucleotide substitution and a strict clock LogNormal(mean=0.001, sigma=0.1) clock rate prior (units: substitutions per site per year).

Introduction (i.e. transition from global to local) and export (local to global) events were assumed to be rare occurrences, and introductions were assumed much more common than exports, reflecting prior knowledge about epidemics originating overseas and then coming to New Zealand. The prior on introductions was Exponential(mean=0.1) migrations per year and Exponential(mean=0.001) for exports. The (constant) effective population sizes of the two demes had independent LogNormal(mean=0.2, sigma=0.4) priors. We sampled the posterior distribution using Markov chain Monte Carlo (MCMC) running each analysis for 100 million steps, sampling states every 10,000 steps, and discarding the first 10% as burn-in. Convergence was assessed in Tracer v1.6^44^, ensuring that all analyses had over 200 effective samples for all reported parameters. All 6/6 RSV-A analyses and 2/6 RSV-B analyses were completed to this standard (the failed analyses are not reported here). The number of introductions of RSV-A and RSV-B over time was estimated using the Babel package in BEAST2 and the results were summarised in R (https://ww.R-Project.org/)^45^.

## Results & Discussion

### The elimination and reintroduction of RSV during the COVID-19 pandemic

In 2021, New Zealand experienced a large epidemic of RSV, following on from a brief period of quarantine-free travel with Australia and other countries in the South Pacific (Figure 1). To better understand the transmission dynamics of RSV before, during, and after this outbreak, we generated RSV genomes from patients in New Zealand over eight years. In total, 1,471 viral genomes, including 756 RSV-A and 715 RSV-B sequences, were generated from New Zealand between 2015 and 2022. Both RSV-A and RSV-B co-circulated each year in relatively even prevalence. Genomes were generated from the male and female sexes in equal proportions, and samples were mostly collected from young children with an overall median age of 1 (range 0 to 99) (Figure 1). Severity of RSV infections can be inferred by the surveillance platform from which the sample originated. Among samples from a known origin, included in this study, 83% were referred from hospital-based surveillance, including outpatients and ICU, meaning that the vast majority of genomes were most likely generated from severe infections (Figure 1).

**Figure 1.**
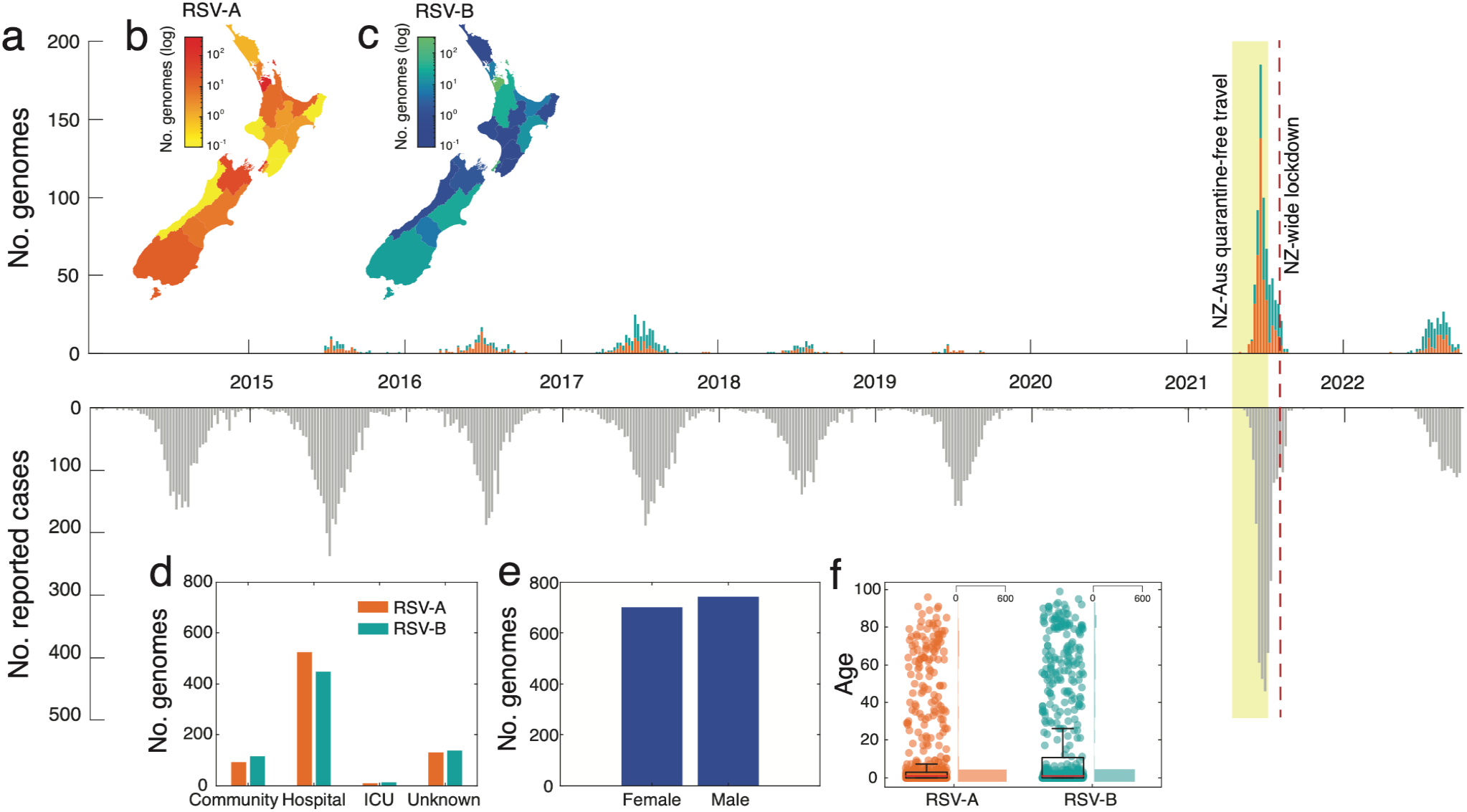
**(a)** Number of RSV-A (orange) and RSV-B (green) genomes generated over time based on sample collection date. The reflected grey bars below show the number of reported RSV positive cases in New Zealand over the same timeframe^46^. **(b-c)** Map of New Zealand, coloured by the number of positive RSV-A and -B cases in each District Health Board. **(d)** The number of RSV genomes generated from cases reported in the community, hospital, intensive care unit (ICU), and those from unknown sources. **(e)** The number of RSV genomes generated from female and male patients. **(f)** The age distribution of patients from which RSV genomes were generated. A box plot indicates the mean (red), lower and upper quartiles (black), and the scatter points show the raw data. An adjacent vertical histogram shows the frequency of patient ages for RSV-A and RSV-B.

Between 2015 and 2019, annual outbreaks of RSV were typically seasonal with cases peaking in late June, coinciding with mid-winter in New Zealand (Figure 1). In 2020, however, reported RSV cases were reduced by 98% and no community transmission of RSV reported as a result of NPI used to control the spread of SARS-CoV-2. Schools and childcare centres were closed during a four-week nationwide lockdown in April 2020, which effectively eliminated SARS-CoV-2 and other respiratory infections from the New Zealand community^14, 47^. The following year, in April 2021, and for the first time since restrictions were placed on the New Zealand border due to the COVID-19 pandemic, people could travel to and from Australia without having to enter managed quarantine at the border. Within weeks of lifting border restrictions there was an exceptionally large RSV epidemic in New Zealand, with a sharp surge in reported cases of both RSV-A and -B (Figure 1).

The 2021 RSV surge affected a wider age distribution compared to previous years (Supplementary Figure 1). Among people who were sampled in this study, we found a 5% increase in infections among 5-18 year olds and a 15% increase in infections among 19-65 year olds compared to previous years (Supplementary Table 3). This surge cannot be explained by weakened immunity due to COVID-19^48^, as there were fewer than 3,000 reported COVID-19 cases, (in a population of ∼five million) before August 2021. The majority of these COVID-19 cases were associated with returning travellers which had limited community transmission^16^. Rather it was more likely the result of waned immunity due to prolonged lack of viral exposure, as seen in other countries^49-51^. However, other factors related to sample collection may have also contributed to the apparent surge. One factor was a change in diagnostic viral testing regimes that were implemented during the COVID-19 pandemic such as the increased use of point of care testing with an expanded respiratory viral diagnostic assay that included RSV, as well as wider testing in the community that included older children and adults. Increased testing not only inflates case counts but can also skew patient age distribution by incorporating less severe cases^52^. Nevertheless, due to the associated rise in hospitalisations^26^ and the return to normal levels in 2022, it is likely that the 2021 epidemic reflected a period of reduced exposure to RSV and an increase in the immunologically naïve population.

Quarantine-free travel with Australia was brought to an abrupt end in July 2021 due to the uncontrolled spread of the SARS-CoV-2 Delta Variant of Concern in Australia^53^. Weeks later, New Zealand entered its second nationwide lockdown due to the incursion of Delta SARS-CoV-2, which was genomically linked to cases in Australia^25^. The lockdown caused the number of reported RSV cases to dramatically reduce (Figure 1).

### Inferring the sources of the 2021 RSV outbreaks using phylodynamics

Phylogenomic analysis of RSV-A and RSV-B genomes, together with genomes sampled from Australia and the rest of the world, showed that in the pre-pandemic phase, genomes sampled from New Zealand were widely distributed among globally sampled genomes, falling among a range of genomic lineages. In 2021, however, genomes sampled from New Zealand formed only a small number of tightly grouped clusters. In particular, RSV-A genomes sampled in 2021 fell into only two distinct lineages (Figure 2). One clade of RSV-A genomes, predominantly sampled from Auckland and surrounding districts, formed a monophyletic clade with no close-in-time sampled genomic ancestors, and unrelated to previously circulating New Zealand lineages (Figure 2). Due to the increased testing regimes coupled with managed quarantine at the border for all arrivals besides those from Australia and the South Pacific, this lineage is unlikely to represent undetected transmission in New Zealand. Rather, it is more likely that this lineage is related to unsampled genomes, most probably in Australia.

**Figure 2.**
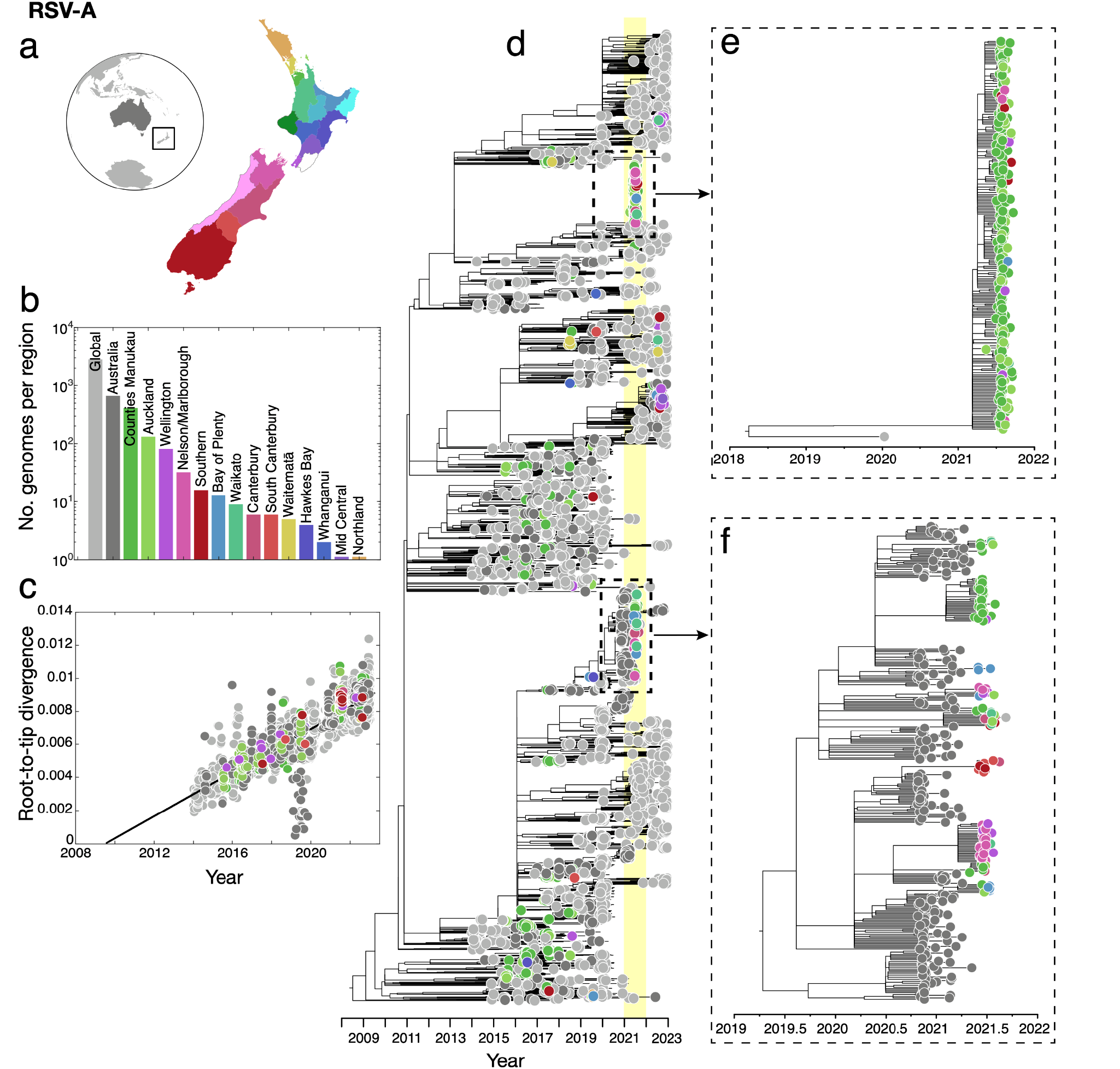
**(a)** Map of New Zealand (where regions are categorised by District Health Board (DHB)), Australia (dark grey), and the rest of the world (light grey). **(b)** Number of RSV-A genomes sampled per region, where colours correspond to those in **(a). (c)** Root-to-tip regression analysis of RSV-A genomes versus sampling time. **(d)** Maximum likelihood time-scaled phylogenetic tree showing 756 RSV-A genomes sampled from New Zealand (coloured circles based on DHB region), 662 RSV-A genomes sampled from Australia (dark grey) and 2,913 RSV-A genomes sampled from the rest of the world (light grey). A yellow vertical bar highlights the year 2021 and a dotted box shows the major New Zealand clades sampled. **(e-f)** Maximum likelihood time-scaled phylogenetic trees showing the major clades sampled in New Zealand during 2021 and their closest sampled genetic relatives.

A second cluster of New Zealand 2021 genomes were grouped with those sampled from Australia during the months prior (Figure 2). This 2021 New Zealand cluster was due to multiple introductions, forming several monophyletic clades among Australian samples, and included genomes from across a wide geographic region including the North and South Islands of New Zealand. Both the highlighted clusters of RSV-A genomes were determined to be the ON1-like genotype based on their G-gene sequences (genomic clade A.3 based on Chen et al.^39^), which has become globally dominant in recent years^54, 55^. This genotype, which evolved from the previous dominant RSV-A genotype NA-1, has a characteristic 72 nucleotide duplication in the G-gene^56^. All RSV-A genomes sampled in New Zealand conformed to a root-to-tip linear regression reflecting a temporal signal in these data (Pearson correlation coefficient, *r* = 0.88, *p*<10^−4^).

Similarly, RSV-B genomes sampled in 2021 were clustered tightly with those sampled in Australia in the months leading up to the outbreak in New Zealand (Figure 3). These genomes formed several large monophyletic clades among Australian samples and largely clustered according to geographic location in New Zealand. We again found a temporal signal in the New Zealand RSV-B data, illustrated by a root-to-tip linear regression (Pearson correlation coefficient, *r* = 0.93, *p*<10^−4^). These genomes were classified as the BA9 genotype based on their G-gene sequences (or the B.5.8 genomic clade^39^). BA9 is one of 15 genotypes within the BA lineage reported since its emergence in around 1998^57^. Genotype BA9 was first identified in Japan in 2006^58^ and, like all BA genotypes, is characterised by a 60 nucleotide duplication in the G-gene^59^. Over the last decade, BA9 has become globally dominant and is now found worldwide^54, 56, 60-62^. Overall, the RSV-A and RSV-B genomes sampled in 2021 were most likely from new introductions into New Zealand rather than from cryptically circulating lineages that remained undetected in 2020. Indeed, our data support that RSV transmission was eliminated in 2020 and both subtypes were reintroduced in 2021, most likely from Australia following the lifting of border restrictions including quarantine.

**Figure 3.**
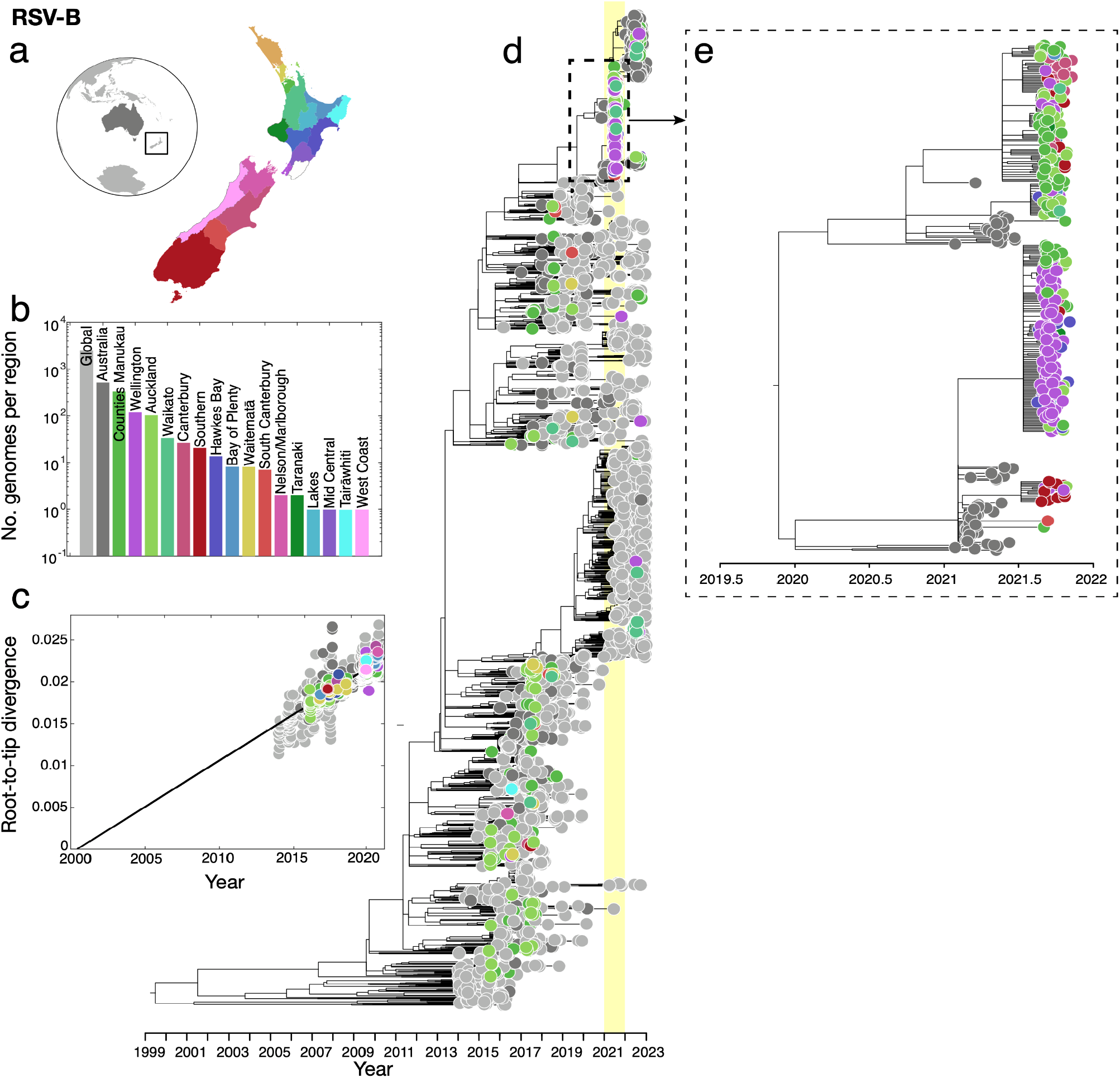
**(a)** Map of New Zealand (where regions are categorised by District Health Board (DHB)), Australia (dark grey), and the rest of the world (light grey). **(b)** Number of RSV-B genomes sampled per region, where colours correspond to those in **(a). (c)** Root-to-tip regression analysis of RSV-B genomes versus sampling time. **(d)** Maximum likelihood time-scaled phylogenetic tree showing 715 RSV-B genomes sampled from New Zealand (coloured circles based on DHB region), 518 RSV-B genomes sampled from Australia (dark grey) and 2,577 RSV-B genomes sampled from the rest of the world (light grey). A yellow vertical bar highlights the year 2021 and a dotted box shows the major New Zealand clade sampled. **(e)** Maximum likelihood time-scaled phylogenetic tree showing the major clades sampled in New Zealand during 2021 and their closest sampled genetic relatives.

### Examining the spatial transmission dynamics of RSV over time

We estimated the spatial transmission dynamics of RSV over time to elucidate how the number of viral introductions changed as a result of NPI, particularly border restrictions, used during the COVID-19 pandemic. While the number of people arriving in New Zealand peaked in early 2020 following an announcement that the border would be closed, a nationwide lockdown meant that any potential new arrivals of RSV did not lead to onward transmission in the community (Figure 4). We inferred the number of introductions of both RSV-A and RSV-B into New Zealand between 2015 and 2022 using genomes sampled from both New Zealand and abroad. Although genomic sequences were too sparse to capture seasonal patterns between 2015 and 2019, introductions of both RSV-A and RSV-B decreased in 2020 resulting in near-zero introductions of either subtype (Figure 4). These results confirm that the number of introductions dramatically increased the following year in 2021, particularly for RSV-A, coinciding with quarantine-free travel to and from Australia. In 2022, RSV-B showed a larger number of introductions, which is consistent with the multiple different lineages that were circulating in 2022. Although both subtypes were introduced many times throughout the eight years (around 80 for RSV-A and 100 for RSV-B), only a small number of these introductions eventuated in large detected outbreaks in 2021 (Figures 2 and 3). This reflects the immunologically naive population of 2021 combined with the highly stochastic nature of disease transmission, which often occurs through superspreader events^63,64^. In contrast, very few export events were identified. Our analysis showed that RSV-B mutated slightly faster than RSV-A (Table 1), and at a comparable rate with previous studies^65^. These experiments proved quite robust to genome subsampling, with each replicate yielding similar clock rates and migration counts.

**Figure 4.**
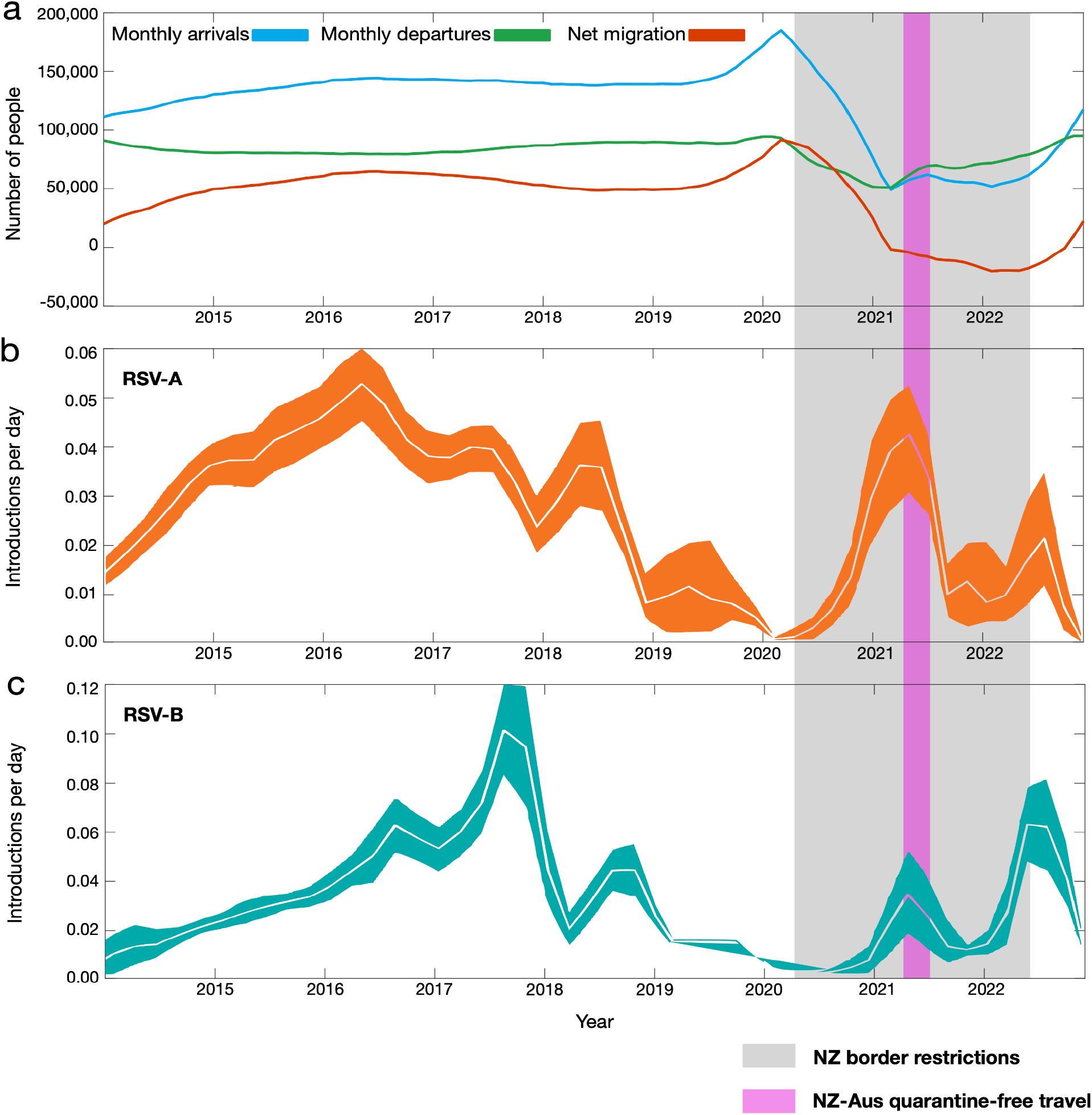
**(a)** Number of people arriving into New Zealand (blue), number of people departing New Zealand (green) and the overall net migration of people into New Zealand (red). (**b-c)** Estimated migration rates of RSV-A and-B into New Zealand over time (mean estimates are shown in white and 95% credible intervals are coloured). A grey shaded area shows the period in which New Zealand underwent border restrictions during the COVID-19 pandemic. A purple shaded area shows the quarantine-free travel period with Australia.

### Genomic diversity of RSV before and after the COVID-19 pandemic

Despite the border-opening associated increase in both viral introductions and genomic surveillance in New Zealand, the surge of introductions in 2021 had very little genomic diversity compared to the pre-pandemic phase as shown by the limited number of New Zealand genomic clusters in our phylogenetic analysis. Similar genomic bottlenecks have been reported in Europe^66^ and Australia^11^, where major lineages of RSV were collapsed following NPI during the COVID-19 pandemic. The resurgence of RSV in 2021 led to very little genomic diversity in circulating lineages, with only two major clades of RSV-A and one clade of RSV-B being reintroduced into New Zealand. Nevertheless, this reduced genetic diversity was seemingly short lived with more widespread lineages reappearing by 2022 when border restrictions were removed. Overall, the extensive NPI used to control SARS-CoV-2 in New Zealand significantly impacted the epidemiology of other respiratory viral infections, including the near-elimination and diminished genetic diversity of typically seasonal viruses.

## Data Availability

The genomes generated are available on GISAID under two blocks of consecutive accession numbers (EPI_ISL_16959469 - EPI_ISL_16960152 and EPI_ISL_19206151 - EPI_ISL_19207488).

https://www.gisaid.org

## Acknowledgements

We gratefully acknowledge all data contributors, i.e. the Authors and their originating laboratories, responsible for obtaining the specimens, and their submitting laboratories for generating the genetic sequence and metadata and sharing via the GISAID Initiative, on which this research is based.

We would like to thank the participants of the SHIVERS WellKiwi cohorts for their continued enrollment and support in our research. Second, we would like to thank the WellKiwi Clinical and Study teams for SHIVERS research, the organisation and collection of participant samples and metadata used in this study. We would also like to thank the clinical virology and genomic sequencing teams at the Institute of Environmental Science and Research for processing, testing and sequencing all of the samples collected.

We would like to acknowledge staff from all of the diagnostic laboratories and hospital laboratories in New Zealand that take the time and effort to collect, aliquot and refer samples to ESR. A special thanks to Nicolas Zacchi from Pathlab in Bay of Plenty, Radhika Nagappan from Auckland City Hospital, Fifi Tse from Middlemore Hospital, and Samantha Hutton from Awanui Laboratory in Wellington for helping collate retrospective data for the sample analysis. Thank you to former ESR staff who contributed to this study.

## Supplementary Information

**Supplementary Figure 1**. The age distribution of patients from which RSV genomes were generated for each year (RSV-A shown in orange and RSV-B shown in green). A box plot indicates the lower and upper quartiles, and the scatter points show the raw data.

**Supplementary Table 1**. GenBank accession numbers and genotypes of G-gene RSV reference sequences.

**Supplementary Table 2**. GISAID accession numbers of global genomes.

**Supplementary Table 3**. Age groups of people from which RSV genomes were derived between 2015 and 2022.

