## Supplementary Table 1 for "Spatial and temporal transmission dynamics of respiratory syncytial virus in New Zealand before and after the COVID-19 pandemic"

**Supplementary Table 1**. GenBank accession numbers and genotypes of G-gene RSV reference sequences.

| **RSV-A** | | **RSV-B** | |
| --- | --- | --- | --- |
| Accession | Genotype | Accession | Genotype |
| AB175815.1 | GA5 | AB161386.1 | JAB1 |
| AB470478.1 | NA1 | AB161387.1 | JAB1 |
| AF065254.1 | GA4 | AB161388.1 | JAB1 |
| AF065255.1 | GA5 | AB161389.1 | JAB1 |
| AF065256.1 | GA2 | AB161390.1 | JAB1 |
| AF065257.1 | GA1 | AB161391.1 | JAB1 |
| AF065407.1 | GA1 | AB161392.1 | JAB1 |
| AF233900.1 | GA2 | AB161395.1 | JAB1 |
| AF233901.1 | GA6 | AB161399.1 | JAB1 |
| AF233902.1 | GA1 | AB175821.1 | BA2 |
| AF233903.1 | GA5 | AB470481.1 | BA7 |
| AF233904.1 | GA7 | AB470482.1 | BA8 |
| AF233905.1 | GA3 | AB603467.1 | BA9 |
| AF233906.1 | GA5 | AB603469.1 | BA9 |
| AF233907.1 | GA7 | AB603470.1 | BA9 |
| AF233909.1 | GA5 | AB603476.1 | BA7 |
| AF233910.1 | GA7 | AB603477.1 | BA7 |
| AF233913.1 | GA3 | AF013254.1 | GB1 |
| AF233914.1 | GA1 | AF065250.1 | GB1 |
| AF233915.1 | GA2 | AF065251.1 | GB2 |
| AF233916.1 | GA5 | AF233924.1 | GB4 |
| AF233917.1 | GA1 | AF233928.1 | GB4 |
| AF233918.1 | GA6 | AF233929.1 | GB3 |
| AF233919.1 | GA5 | AF233931.1 | GB4 |
| AF233920.1 | GA3 | AF233932.1 | GB3 |
| AF233921.1 | GA3 | AF309676.1 | SAB2 |
| AF233923.1 | GA2 | AF309678.1 | SAB2 |
| AF348803.1 | GA5 | AF348811.1 | SAB3 |
| AF348804.1 | GA7 | AF348812.1 | SAB3 |
| AF348807.1 | SAA1 | AF348813.1 | SAB3 |
| AF348808.1 | SAA1 | AF348817.1 | GB3 |
| AF448498.1 | GA2 | AF348821.1 | SAB2 |
| AY114149.1 | GA2 | AF348824.1 | GB4 |
| AY114150.1 | GA5 | AF348825.1 | SAB1 |
| AY114151.1 | GA2 | AY333361.1 | URU2 |
| AY146435.1 | GA2 | AY333364.1 | BA1 |
| AY146437.1 | GA5 | AY488804.1 | URU1 |
| AY472086.1 | GA2 | AY488805.1 | URU1 |
| AY472094.1 | GA5 | AY524573.1 | SAB1 |
| AY911262.1 | PRO | AY660682.1 | SAB1 |
| JF920069.1 | GA1 | AY672691.1 | GB4 |
| JN257694.1 | ON1 | AY672698.1 | GB4 |
| JX256960.1 | NA2 | AY751087.1 | BA7 |
| KC297260.1 | NA3 | AY751105.1 | BA6 |
| KC297277.1 | NA3 | AY751111.1 | BA6 |
| KC297292.1 | NA3 | AY751116.1 | BA6 |
| KC297324.1 | NA4 | AY751117.1 | BA6 |
| KC297381.1 | NA4 | AY751119.1 | BA2 |
| KF300972.1 | NA1 | AY751121.1 | BA2 |
| KF300973.1 | ON1 | AY751122.1 | BA2 |
| KP792358.1 | NA1 | AY751123.1 | BA2 |
| KP792359.1 | NA1 | AY751174.1 | GB12 |
| KP792361.1 | ON1 | AY751237.1 | GB6 |
| KP792362.1 | ON1 | AY751239.1 | GB6 |
| KP792365.1 | ON1 | AY751241.1 | GB6 |
| KP792370.1 | ON1 | AY751256.1 | GB1 |
| KP792373.1 | ON1 | AY751280.1 | GB5 |
| KP792374.1 | ON1 | AY751281.1 | GB5 |
| KP792375.1 | ON1 | DQ171841.1 | NZB2 |
| M11486.1 | GA1 | DQ171842.1 | NZB2 |
| M17212.1 | PRO | DQ171843.1 | NZB2 |
| Z33414.1 | GA3 | DQ171844.1 | NZB2 |
| Z33416.1 | GA3 | DQ171845.1 | NZB2 |
| Z33417.1 | GA7 | DQ171846.1 | NZB2 |
| Z33422.1 | GA2 | DQ171847 | NZB2 |
| Z33426.1 | GA3 | DQ171849.1 | GB2 |
| Z33427.1 | GA1 | DQ171858.1 | GB2 |
| Z33430.1 | GA5 | DQ171863.1 | NZB1 |
| Z33431.1 | GA1 | DQ171865.1 | NZB1 |
| Z33432.1 | GA1 | DQ171867.1 | GB13 |
| Z33455.1 | GA7 | DQ171878.1 | GB13 |
| Z33494.1 | GA5 | DQ227363.1 | BA1 |
|  |  | DQ227364.1 | BA1 |
|  |  | DQ227368.1 | BA1 |
|  |  | DQ227370.1 | BA3 |
|  |  | DQ227373.1 | BA1 |
|  |  | DQ227374.1 | BA1 |
|  |  | DQ227375.1 | BA3 |
|  |  | DQ227377.1 | BA2 |
|  |  | DQ227389.1 | BA2 |
|  |  | DQ227393.1 | BA2 |
|  |  | DQ227396.1 | BA4 |
|  |  | DQ227397.1 | BA3 |
|  |  | DQ227403.1 | BA3 |
|  |  | DQ227408.1 | BA4 |
|  |  | EU635867.1 | BA9 |
|  |  | HM459858.1 | BA4 |
|  |  | HM459864.1 | BA7 |
|  |  | HM459865.1 | BA7 |
|  |  | HM459866.1 | BA7 |
|  |  | HM459867.1 | BA7 |
|  |  | HM459868.1 | BA7 |
|  |  | HM459870.1 | BA7 |
|  |  | HM459871.1 | BA8 |
|  |  | HM459872.1 | BA8 |
|  |  | HM459875.1 | BA8 |
|  |  | HM459878.1 | BA9 |
|  |  | HM459880.1 | BA9 |
|  |  | HM459881.1 | BA9 |
|  |  | HM459882.1 | BA9 |
|  |  | HM459883.1 | BA10 |
|  |  | HM459884.1 | BA10 |
|  |  | HM459886.1 | BA10 |
|  |  | HM459888.1 | BA10 |
|  |  | HM459890.1 | BA10 |
|  |  | HM459891.1 | BA10 |
|  |  | JN119979.1 | SAB4 |
|  |  | JN119987.1 | SAB4 |
|  |  | JN119989.1 | SAB4 |
|  |  | JN120007.1 | SAB4 |
|  |  | JX256976.1 | BA12 |
|  |  | JX256977.1 | BA12 |
|  |  | KC297426.1 | BA10 |
|  |  | KC297428.1 | CB1 |
|  |  | KC297456.1 | BA-C |
|  |  | KC297471.1 | THB |
|  |  | KC297486.1 | BA-C |
|  |  | KF246585.1 | BA12 |
|  |  | KF246586.1 | BA12 |
|  |  | KF246607.1 | BA9 |
|  |  | KF246624.1 | BA9 |
|  |  | KF246629.1 | BA9 |
|  |  | KF300952.2 | BA14 |
|  |  | KF300953.2 | BA14 |
|  |  | KF300954.2 | BA14 |
|  |  | KF300955.2 | BA14 |
|  |  | KF300957.2 | BA14 |
|  |  | KF300958.2 | BA14 |
|  |  | KF300959.2 | BA14 |
|  |  | KF300960.2 | BA14 |
|  |  | KF300970.2 | BA14 |
|  |  | KP336523.1 | BA11 |
|  |  | KP336524.1 | BA11 |
|  |  | KP336525.1 | BA11 |
|  |  | KP336526.1 | BA11 |
|  |  | KP336527.1 | BA11 |
|  |  | KP336528.1 | BA11 |
|  |  | KP336530.1 | BA11 |
|  |  | KP336531.1 | BA11 |
|  |  | KP336532.1 | BA11 |
|  |  | KP336533.1 | BA11 |
|  |  | KP336534.1 | BA11 |
|  |  | KP336535.1 | BA11 |
|  |  | KP336536.1 | BA11 |
|  |  | KP336537.1 | BA11 |
|  |  | KP336538.1 | BA11 |
|  |  | KP336540.1 | BA11 |
|  |  | KP336541.1 | BA11 |
|  |  | KP336542.1 | BA11 |
|  |  | KP336543.1 | BA11 |
|  |  | KP336544.1 | BA11 |
|  |  | KP336545.1 | BA11 |
|  |  | KU254638.1 | BA-CCB |
|  |  | KU254641.1 | BA-CCA |
|  |  | KU254642.1 | BA-CCA |
|  |  | KU254643.1 | BA-CCB |
|  |  | KX262619.1 | BA13 |
|  |  | KX262621.1 | BA13 |
|  |  | KX262625.1 | BA13 |
|  |  | KX262638.1 | BA13 |
|  |  | KX371866.1 | BA14 |
|  |  | KX371867.1 | BA14 |
|  |  | KX371868.1 | BA14 |
|  |  | M17213.1 | PRO |
|  |  | M73540.1 | GB1 |
|  |  | M73541.1 | GB1 |
|  |  | M73542.1 | GB1 |
