## Supplementary Table 3 for "Spatial and temporal transmission dynamics of respiratory syncytial virus in New Zealand before and after the COVID-19 pandemic"

**Supplementary Table 2**. Age groups of people from which RSV genomes were derived between 2015 and 2022.

| **Age** | **2015** | **2016** | **2017** | **2018** | **2019** | **2020** | **2021** | **2022** |
| --- | --- | --- | --- | --- | --- | --- | --- | --- |
| Under 5 | 52 | 109 | 177 | 39 | 8 | 0 | 474 | 261 |
| Between 5 and 18 | 2 | 2 | 3 | 5 | 3 | 0 | 31 | 10 |
| Between 19 and 65 | 4 | 3 | 11 | 7 | 9 | 0 | 106 | 31 |
| Over 65 | 6 | 6 | 19 | 2 | 3 | 0 | 79 | 9 |
