## Supplementary figures and images for "Spatial and temporal transmission dynamics of respiratory syncytial virus in New Zealand before and after the COVID-19 pandemic"

### Supplementary Figure 1

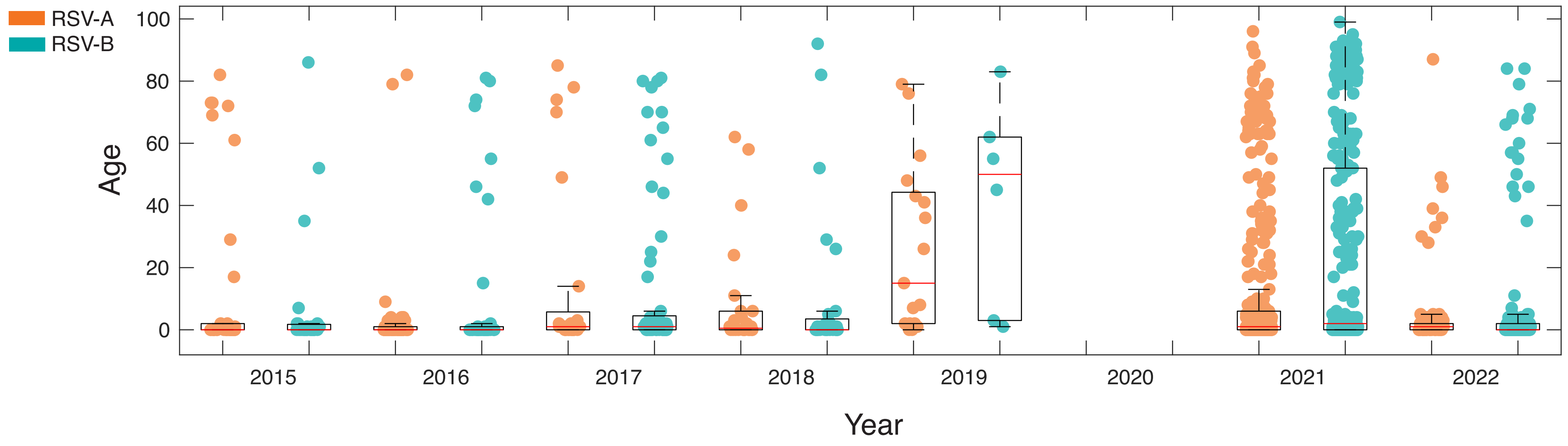
